# Intranasal oxytocin enhances approach-related EEG frontal alpha asymmetry during engagement of direct eye contact

**DOI:** 10.1101/2020.02.26.20028266

**Authors:** Javier R. Soriano, Nicky Daniels, Jellina Prinsen, Kaat Alaerts

## Abstract

The neuropeptide oxytocin is suggested to exert a pivotal role in a variety of complex human behaviors, including interpersonal bonding, trust, and attachment. Recent theories have suggested that the role oxytocin plays in these complex social behaviors involves a modulation of motivational tendencies of approach/avoidance-related behaviors. However, to date, direct neurophysiological evidence supporting this notion is limited. In this double-blind, randomized, placebo-controlled study with parallel design, we assessed the effects of administered intranasal oxytocin in 40 adult men on gaze behavior and a neural marker of approach/avoidance motivational tendencies. Specifically, electroencephalography recordings were performed during the engagement of eye contact with a live model in a naturalistic two-person social context and electroencephalographic frontal alpha asymmetry, an established neurophysiological index of motivational tendencies for approach/avoidance-related behaviors, was assessed. Compared to placebo, a single dose of oxytocin (24 international units) was shown to increase relative left-sided frontal asymmetry upon direct eye contact with a live model, which is indicative of an increase in approach-related motivational tendencies toward the presented eye contact stimulus. Notably, the treatment effect was most prominently observed in participants with lower self-reported social motivation (higher Motivation subscale scores on the Social Responsiveness Scale), indicating that participants with lower social motivation benefitted the most from the administered oxytocin. No treatment-specific changes were identified in terms of gaze behavior toward the eye region of the live model. Together, these observations add neurophysiological evidence to the hypothesized role of oxytocin in modulating approach/avoidance related tendencies and suggest that inter-individual variance in person-dependent factors need to be considered in order to evaluate the potential benefit of intranasal oxytocin as a treatment. This notion is of particular relevance to the variety of neuropsychiatric populations such as autism spectrum disorder, social anxiety disorder and depression, for which intranasal oxytocin is increasingly considered a potential treatment.

## Introduction

The neuropeptide oxytocin is synthesized in the magnocellular neurons of the supraoptic and paraventricular nuclei of the hypothalamus and released peripherally and centrally to act as a hormone and as a neurotransmitter respectively. Amongst its main effects in human physiology, oxytocin is well known for aiding parturition (Blanks & Thornton, 2003) and lactation (Crowley & Armstrong, 1992). In addition, oxytocin has gained increasing interest in the fields of social psychology and neuroscience, whereby it is studied as a neuromodulator that affects social cognition and behavior (Heinrichs *et al*., 2009; Guastella & MacLeod, 2012), and specifically prosocial behaviors (for a detailed review see MacDonald & MacDonald, 2010) such as inter-personal bonding, trust (Kosfeld *et al*., 2005; Baumgartner *et al*., 2008) and generosity (Zak *et al*., 2007).

The Social Approach/Withdrawal hypothesis (Kemp & Guastella, 2011) aims at explaining oxytocin’s complex role in modulating prosocial behavior, suggesting that it induces both a down-regulation of withdrawal (also termed avoidance)-related behavior and an up-regulation of approach-related behavior, by inducing a decrease in the experience of social threat (anxiolytic effect) and an increase in the recognition of social reward-driven stimuli (enhancement of social salience), respectively. Several lines of evidence provide support for oxytocin’s impact on increasing social salience (see Shamay-Tsoory & Abu-Akel, 2016 for a review), by for example, showing increased dwelling time towards the eye region of faces after intranasal administration of oxytocin (Guastella *et al*., 2008; Gamer *et al*., 2010; Auyeung *et al*., 2015). With respect to its anxiolytic effects, studies have found intranasal oxytocin to diminish individual reports of anxiety, as well as cortisol stress responses (Heinrichs *et al*., 2003; Meinlschmidt & Heim, 2007; Riem *et al*., 2020). Additionally, several studies have reported reductions in skin conductance levels, an autonomic measure indicative of physiological arousal, after intranasal oxytocin during stress-inducing tasks (Pitman *et al*., 1993; De Oliveira *et al*., 2012).

However, fewer studies have addressed intranasal oxytocin’s impact on modulating approach/avoidance motivational tendencies directly. In one study by Radke *et al*. (2013), the effect of intranasal oxytocin on approach/avoidance tendencies towards social stimuli was explored by allowing participants to pull a joystick toward them (reflecting approach) or push it away from themselves (indicating avoidance), while observing happy and angry faces with direct or averted gaze. While participants of the placebo group showed typical motivational tendencies (increased approach/avoidance upon happy/angry faces), participants of the oxytocin group showed a general moderation of these tendencies, with even an increased inclination to approach angry faces with direct gaze. The authors argued that intranasal oxytocin facilitated approach in humans in response to social threat, thereby supporting its anxiolytic potential. In a similar study by Theodoridou *et al*. (2013), participants administered with intranasal oxytocin could increase or decrease the stimuli size of pictures displaying facial emotional expressions (social condition) or natural scenes (non-social condition), by pulling a lever towards (approach) or away from them (avoidance) respectively, but no significant effects on approach/avoidance tendencies were identified. Here, the authors argued that the adopted pictures may have lacked enough social animation to elicit social feelings of negative affect and that an effect of intranasal oxytocin may have been more pronounced in more interactive, real-life contexts.

Approach/avoidance motivation systems have been widely studied from a neurophysiological perspective employing the so-called electroencephalographic (EEG) frontal alpha asymmetry index (Gable & Harmon-Jones, 2008; Harmon-Jones & Gable, 2009; Schöne *et al*., 2016), one of the core approach/avoidance neural bases (Harmon-Jones, 2011). This measure indexes relative interhemispheric frontal cortical activity and is usually computed subtracting the alpha power activity of the left fontal hemisphere (measured at electrode F3) from that of the right frontal hemisphere (measured at electrode F4). Since alpha power is widely assumed to reflect cortical inhibition (Jensen & Mazaheri, 2010), a positive asymmetric value, indexing greater relative alpha power in the right frontal hemisphere, implies more inhibition in this region than in its left counterpart. Several studies have shown that greater relative left frontal activity (positive asymmetry score) is linked to trait approach motivation (Harmon-Jones & Allen, 1997; Sutton & Davidson, 1997; Coan & Allen, 2003; Amodio *et al*., 2008), approach motivation to appetitive stimuli (Gable & Harmon-Jones, 2008; Schöne *et al*., 2016), different degrees of approach motivation irrespective of positive affect (Harmon-Jones *et al*., 2008*a*), and approach motivation depending on the perceived degree of choice (Harmon-Jones *et al*., 2008*b*; Harmon-Jones *et al*., 2011*a*). Despite the notion that increasing approach motivation may be one of the core mechanisms through which intranasal oxytocin mediates its prosocial effects (Kemp & Guastella, 2011), there have been to date no studies directly assessing the effect of intranasal oxytocin on EEG frontal alpha asymmetry.

In the present study, a double-blind, randomized, placebo-controlled, parallel design was adopted to examine the effect of a single dose of intranasal oxytocin on EEG frontal alpha asymmetry upon presentation of direct eye contact, which entails a highly salient social cue. Considering prior concerns that the effects of intranasal oxytocin may be more pronounced in interactive, real-life contexts (Theodoridou *et al*., 2013), the eye contact cues were conveyed by a real live model within a naturalistic two-person social context.

In prior studies with neurotypical adults (Hietanen *et al*., 2008) and children (Kylliäinen *et al*., 2012), direct gaze conveyed by a live model has been shown to form a salient cue for eliciting a relative left-sided frontal activation (lower alpha power recorded over the left, compared to the right frontal electrode), associated with approach motivation, while averted gaze or closed eyes elicited a relative right-sided frontal activation (higher alpha power recorded over the left, compared to the right frontal electrode) associated with withdrawal motivation. Importantly, this pattern was not revealed in young children with autism spectrum disorder (Kylliäinen *et al*., 2012; Lauttia *et al*., 2019) and it was decreased in individuals with high social anxiety (Myllyneva *et al*., 2015), indicating that in these patient populations, another person’s direct gaze does not elicit the expected approach-motivation related brain activity.

In line with the social approach/avoidance account of oxytocin, we generally hypothesized a single dose of intranasal oxytocin to induce a relative increase in left-sided frontal asymmetry (associated with approach motivation) upon the presentation of live direct eye gaze. Furthermore, taking into account that person-dependent factors can modulate treatment effects of intranasal oxytocin (Bartz *et al*., 2011), we included the Social Responsiveness Scale (SRS; Constantino *et al*., 2003) in its self-reported version to assess social proficiency.

Consistently with prior observations of a lack of approach-related brain activity in autism spectrum disorder children and individuals with high social anxiety, we generally hypothesized the effects of intranasal oxytocin on EEG frontal alpha asymmetry to be most pronounced in participants with low baseline social responsiveness.

## Materials and methods

### Participants

A total of 56 healthy adults (all right-handed, Dutch-speaking men) were recruited to participate in this double-blind, randomized, placebo-controlled study with parallel design.

Due to technical difficulties, electroencephalography (EEG) and eye tracking recordings were not available for six participants (3 oxytocin and 3 placebo). Additionally, EEG data were not available from 10 additional participants (8 oxytocin and 2 placebo) due to excessive signal artifacts. Thus, EEG analyses were performed for a total of 40 participants (17 oxytocin, mean age 22.0 ± 3.0 years; 23 placebo, 22.65 ± 3.33 years). Only male participants were included to avoid the potential confound of gender-dependent effects of oxytocin (Zink & Meyer-Lindenberg, 2012). Participants abstained from alcohol and caffeine 24 hours before testing. Written informed consent was obtained from all participants prior to the study. Consent forms and study design were approved by the Ethics Committee Research UZ / KU Leuven in accordance to The Code of Ethics of the World Medical Association (Declaration of Helsinki). The study procedure was pre-registered at ClinicalTrials.gov (ClinicalTrials.gov Identifier: NCT03272321).

Note that participants were recruited to participate in a larger study, including additional physiological recordings of respiration, cardiac activity and skin conductance. A detailed report of the effects of intranasal oxytocin on skin conductance is provided elsewhere (Daniels *et al*., 2020). Overall, the procedures outlined below for nasal spray administration and eye gaze stimuli are the same as those described in (Daniels *et al*., 2020).

### Nasal spray administration

Participants were randomly assigned to receive a single dose of oxytocin or placebo based on a computer-generated randomized order. Except for the manager of randomization and masking of drug administration, all participants and research staff conducting the trial were blind to treatment allocation. In correspondence with previous studies (see Guastella & MacLeod, 2012 for a review), participants received 24 international units (IU) of oxytocin (Syntocinon®, Sigma-tau) or placebo containing a saline natriumchloride solution. Participants received three puffs per nostril in an alternating fashion with each puff containing 4 IU. Participants were asked to first remove air present in the nasal spray by pumping the spray until a fine mist was observed. For inhalation of the spray, they were instructed to take a deep breath through the nose and to tilt their head slightly backwards during nasal administration in order to minimize gravitational loss of the spray. All participants were monitored onsite until approximately one hour after nasal spray administration and were then screened for potential side effects, and administered the 32-item short version questionnaire of the Profile of Mood States (POMS; Mcnair & Lorr, 1964) for potential changes in mood states (for a detailed report see Daniels *et al*., 2020, and **Supplementary Tables 1 and 2**). In short, only minimal, non-treatment specific side effects were reported, although note that a larger proportion of participants receiving the oxytocin spray reported a (mild) headache (oxytocin group: 6 out of 28; placebo group: 1 out of 27). At the end of the experimental session, participants were asked if they thought they had received oxytocin or placebo, or whether they were uncertain about the received spray. The majority of participants reported to be uncertain about the received spray (oxytocin: 48%; placebo: 48%) or thought they had received the placebo nasal spray (oxytocin: 32%; placebo: 40%). The proportion of participants who believed that they had received the oxytocin treatment was not significantly larger in the actual oxytocin group (20%), compared to the placebo group (12%) (Pearson Chi-square = 0.60; *p*= 0.44).

**Table 1.** Pre-to-post treatment changes in main outcome variables. Mean pre-to-post change scores are listed separately for each treatment group (oxytocin, placebo). *T* and *p* values correspond to between-group differences in pre-to-post change scores (independent t-test).

|  | Oxytocin |  | Placebo |  | Group difference |  | Effect size |
| --- | --- | --- | --- | --- | --- | --- | --- |
|  | *N* | *Mean ± SD* | *N* | *Mean ± SD* | *t* | *p* | *Cohen's d* |
| **EEG Frontal Alpha Asymmetry Change** |  |  |  |  |  |  |  |
| Eyes open | 17 | 0.15 ± 0.40 | 23 | -0.07 ± 0.28 | **2.069** | **0.04\*** | **0.64** |
| Eyes closed | 17 | 0.11 ± 0.39 | 23 | -0.05 ± 0.34 | 1.443 | 0.16 | 0.46 |
| **Gaze behavior Change** |  |  |  |  |  |  |  |
| Fixation count eyes open | 25 | -3,77 ± 30,46 | 25 | -3,66 ± 16,24 | 0.02 | 0.99 | 0 |
| Fixation count eyes closed | 25 | -3,18 ± 27,78 | 25 | -3,78 ± 15,50 | -0.09 | 0.93 | 0.02 |
| Fixation time eyes open | 25 | -3,97 ± 30,03 | 25 | -2,35 ± 16,04 | 0.24 | 0.81 | 0.07 |
| Fixation time eyes closed | 25 | -4,41 ± 28,79 | 25 | -3,80 ± 15,29 | 0.09 | 0.93 | 0.03 |

\* *p* < .05. SD: ± standard deviation.

### Experimental procedure and stimuli

Participants were seated in front of a 20 × 30 cm voltage-sensitive liquid crystal shutter screen (DreamGlass Group, Spain) attached to a black frame separating the participant from the live model (similar setup as used by Prinsen *et al*., 2019). Participants were instructed to observe and pay close attention to the stimuli presented through the screen. The presented stimuli were one of two live female models, of which only the face was visible, while sitting with eyes open and gazing towards the participant or sitting with eyes closed. The models were trained to act similarly towards participants, were unknown to them and only had a brief standardized interaction with them before stimuli presentation. For each participant, the same model was presented pre- and post-nasal spray administration. Participants were seated 60 cm in front of the shutter, with their head at the same height as the model’s face, with an overall distance of 110 cm between the participant and the model. Nexus Trigger Interface in combination with E-Prime software was used to trigger the liquid crystal window to shift from an opaque to a transparent state. Each stimulus condition (model with eyes open or eyes closed) was presented 10 times with a duration of 5 sec each in a semi-random fashion (no more than three consecutive trials of the same type). The inter-stimulus interval (ISI) varied with a minimum of 20 seconds. During all trials, electroencephalography recordings and eye tracking measures were acquired. Additionally, skin conductance, blood volume pulse and respiration measures were also acquired, but are not part of the current report (see Daniels *et al*., 2020 for a report on skin conductance).

### Assessment of person-dependent factors

Prior to neurophysiological assessments and nasal spray administrations, participants completed the Social Responsiveness Scale (SRS; Constantino *et al*., 2003). The 64-item Dutch adult self-report version of the SRS comprises four subscales examining social communication, social awareness, social motivation and rigidity/repetitiveness, using a four-point Likert-scale.

### Electroencephalography (EEG) recordings and data handling

The Nexus-32 multimodal acquisition system and BioTrace+ software (version 2015a, Mind Media, The Netherlands) were used to collect electroencephalography recordings. Continuous EEG was recorded with a 21-electrode cap (MediFactory, The Netherlands) positioned according to the 10–20 system. Skin abrasion and electrode paste (Nuprep) were used to reduce electrode impedance. Vertical (VEOG) and horizontal (HEOG) eye movements were recorded by placing pre-gelled foam electrodes (Kendall, Germany) above and below the left eye (VEOG) and next to the left and right eye (HEOG). The sampling rate of the VEOG and HEOG recordings was 2048 Hz. The EEG signal was amplified using a unipolar amplifier and referenced offline to linked mastoids. The sampling rate for the digitized EEG signal was 256 Hz. The EEG signal was filtered using a 0.5 Hz low cut-off, a 70 Hz high cut-off and 50 Hz notch filter. After visual inspection, deflections resulting from eye blinks and horizontal eye movements (assessed using the VEOG and HEOG channels), were removed by the implemented Independent Component Analysis (ICA) module in BrainVision Analyzer 2 (Jung *et al*., 2000). Thereafter, the 5-s stimulus periods were segmented into epochs of 1 s with an overlap of 0.5 s, resulting in 90 epochs for each condition (eyes open, eyes closed). Segments with residual artifacts exceeding ± 100 µV in amplitude were rejected.

Averaged power spectra were calculated over all epochs and trials separately for each condition (eyes open, eyes closed), and power spectral density (µV^2^/Hz) within the alpha band (8-13 Hz) was computed and ln-transformed. For each condition, frontal electrode pairs F3/F4 were used to calculate the frontal alpha asymmetry score, by subtracting the ln-transformed alpha power values of the left hemisphere (F3) from that of the right hemisphere (F4); [ln(alpha power F4) – ln(alpha power F3)]. A positive asymmetry value indexes stronger relative left-sided frontal brain activity (lower frontal alpha in left F3, compared to right F4) associated with approach motivation, whereas a negative value indexes stronger relative right-sided frontal brain activity (lower frontal alpha in right F4, compared to left F3) associated with avoidance motivation.

### Eye tracking and data handling

Eye tracking data were recorded with a 30 Hz sampling rate using iView and analyzed with BeGaze software (SensoMotoric Instruments; SMI, Germany). The SMI eye tracking glasses were adjusted to the participant’s comfort and 3-point calibrations were performed at regular time intervals. An area of interest (AOI) was specified over the top half of the model’s face to assess, for each condition, the number of fixations that were directed toward the eye region of the model’s face (% fixation count) and the total fixation time spent looking at the eye region of the model’s face (% fixation time).

## Statistical analyses

### Effect of treatment

Pre-to-post treatment changes in EEG frontal asymmetry scores; [ln(alpha power F4post) – ln(alpha power F3post)] – [ln(alpha power F4pre) – ln(alpha power F3pre)], and eye tracking measures [(fixation count post – fixation count pre) and (fixation time post – fixation time pre)] were calculated for all participants, averaged per treatment condition, and subjected to t-tests for independent samples (two-sided) with the categorical factor ‘treatment’ (oxytocin or placebo) to assess treatment-specific effects. Where relevant, Cohen’s *d* effect sizes are reported (calculated as follows: (pre-to-post difference score_oxytocin_ – pre-to-post difference score_placebo_)/pooled standard deviation), where 0.2 is indicative of a small effect, 0.5 a medium effect and 0.8 a large effect (Cohen, 2013).

### Modulation of treatment effect by person-dependent factors

Pearson’s *r* correlation analyses were performed to assess whether the effects of intranasal oxytocin on EEG frontal alpha asymmetry were more pronounced in participants with low baseline social proficiency (SRS). All statistical analyses were executed with SPSS 26 (IBM, USA), and significance levels set at *p* < .05.

### Data availability

The data set discussed in this manuscript will be made available upon reasonable request.

## Results

### Effect of intranasal oxytocin on frontal asymmetry

During the engagement of direct eye contact, a significant effect of treatment was identified (*t*(38)= 2.07; *p*= 0.04; Cohen’s *d*= 0.64, medium size effect), indicating a pre-to-post increase in left-sided frontal asymmetry (associated with approach motivation) in the intranasal oxytocin group, compared to the placebo group (**Fig. 1 and Table 1**). No significant effect of treatment was evident during the eyes closed condition (*t*(38)= 1.44; *p*= 0.16; Cohen’s *d*= 0.46) (**Fig. 1 and Table 1**).

**Figure 1.**
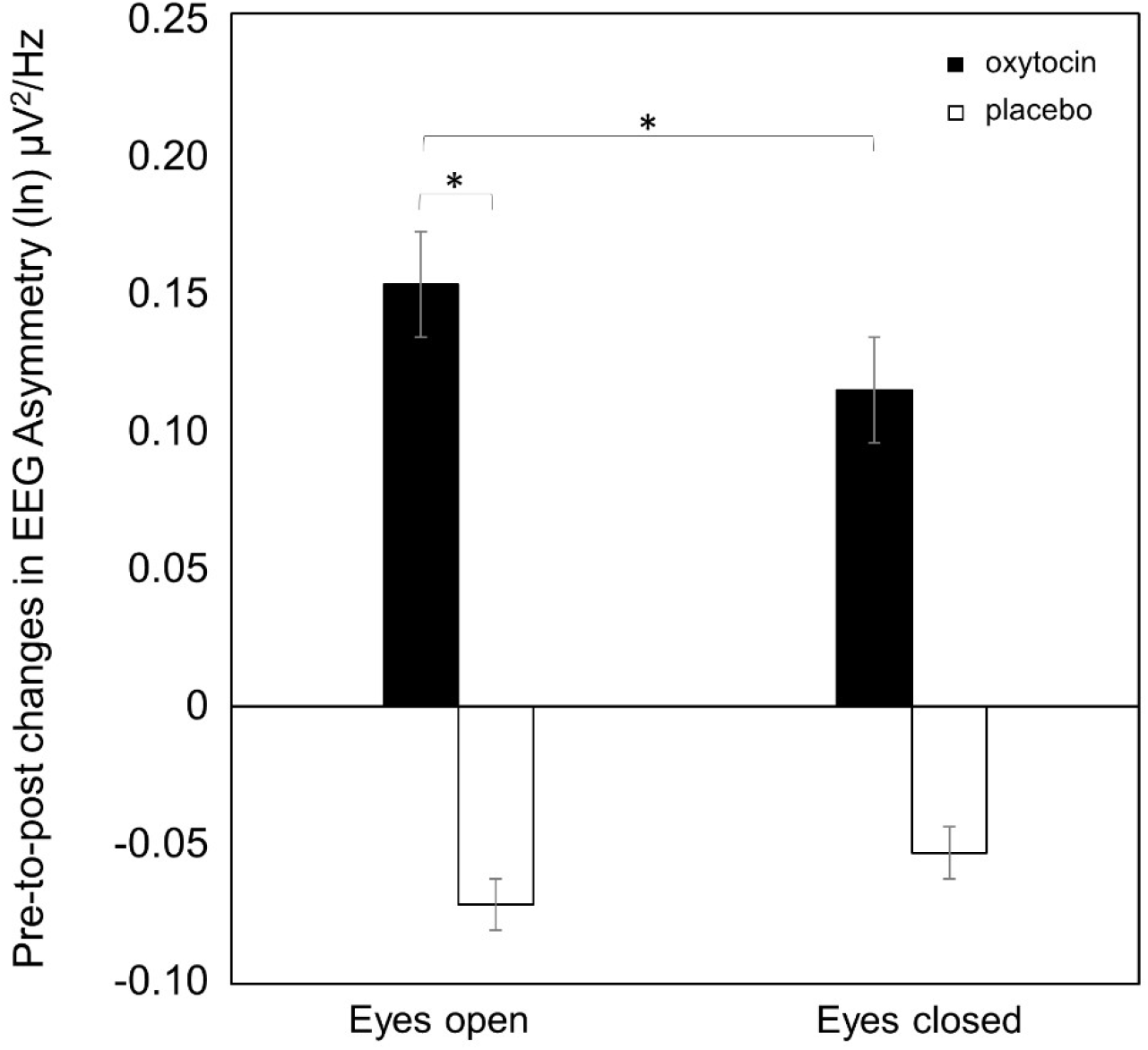
Effect of oxytocin on EEG frontal asymmetry. For each treatment group (oxytocin in black and placebo in white), pre-to-post changes in EEG frontal asymmetry ([F4post-F3post] – [F4pre-F3pre]), are visualized separately for each condition (eyes open, eyes closed). Positive changes in asymmetry score indicate a change towards approach-related motivational tendencies, whereas negative changes in asymmetry scores indicate a change towards less approach-related, or more avoidance-related motivational tendencies. Vertical bars denote +/- standard deviations. *\*p*< .05.

Within-group analyses further confirmed this pattern of results, indicating that in the intranasal oxytocin group, pre-to-post increases in relative left-sided frontal asymmetry (towards approach-motivation) were significantly larger upon the direct eye contact condition, compared to the eyes closed condition (paired-sample t-test: *t*(16)= 2.11; *p* =0.05), whereas in the placebo group, no such effect was revealed (*t*(22)= -0.46; *p*= 0.65).

Contrary to our expectations based on prior work (Hietanen *et al*., 2008), frontal alpha asymmetry scores, investigated separately at pre or post treatment, were not found to be significantly higher upon the presentation of the direct eye contact condition, compared to the eyes closed condition, either in the intranasal oxytocin or placebo group (oxytocin pre: *t*(16) = -0.16 *p* = 0.87; placebo pre: *t*(22) = -0.05 *p* = 0.96; oxytocin post: *t*(16) = 1.28 *p* = 0.22; placebo post: *t*(22) = -0.90; *p* = 0.38).

### Effect of intranasal oxytocin on eye gaze behavior

Administration of intranasal oxytocin (versus placebo) did not significantly modulate the number of fixations directed toward the eye region of the model’s face (% total fixation count) nor the fixation time spent looking at the eye region of the face (% total fixation time), either for the direct eye contact open (fixation count: t(48) = 0.02; *p*=0.99; Cohen’s *d* = 0 ; fixation time: *t*(48) = 0.24; *p=0*.*81*; Cohen’s *d* = 0.07) or eyes closed condition (fixation count: *t*(48) = -0.09; *p=0*.*9* ; Cohen’s *d* = 0.02 ; fixation time: *t*(48) = 0.09; *p=0*.*93* ; Cohen’s *d* = 0.03) (see **Table 1**).

### Modulation of treatment effects by person-dependent factors

The effect of intranasal oxytocin treatment on frontal asymmetry upon the engagement of direct eye contact was modulated by person-dependent factors, indicating that participants of the oxytocin group with higher self-reported scores on the ‘Motivation’ subscale of the Social Responsiveness Scale (SRS), reflecting lower social motivation, showed a stronger treatment-induced increase in relative left-sided frontal asymmetry (increasing approach-motivation) when perceiving direct eye gaze from the model (Pearson *r* = 0.55; *p* = 0.02) (**Fig. 2**). No significant relationship between the SRS Motivation subscale and changes in frontal alpha asymmetry were identified for the placebo group (Pearson *r* = -0.14; *p* = 0.52). The treatment effect on frontal asymmetry was also not significantly modulated by inter-individual variance in the other three SRS subscales (all tests yielding *p* > 0.1).

**Figure 2.**
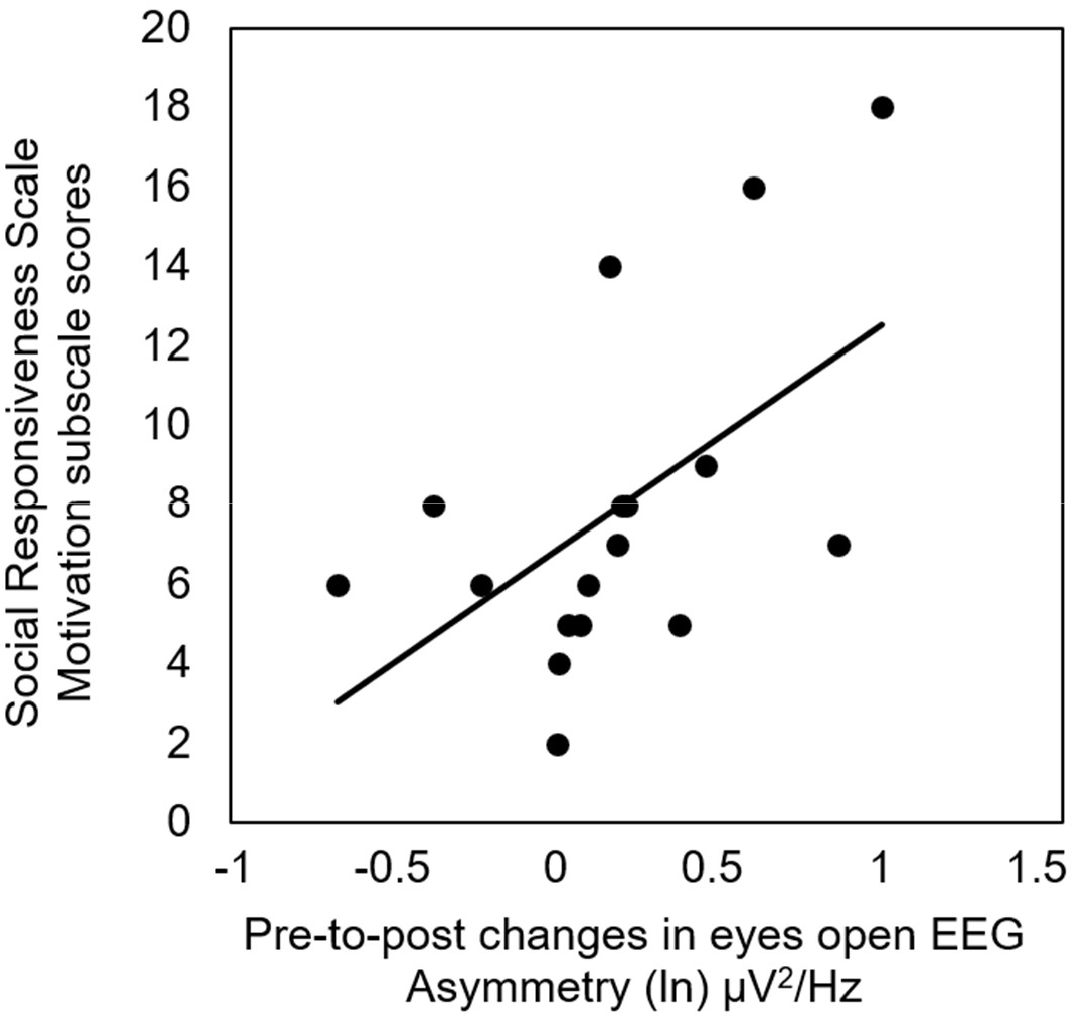
Modulation of frontal asymmetry treatment-induced changes by person-dependent factors in the oxytocin group. Visualization of the relationship between scores on the Motivation subscale of the Social Responsiveness Scale (SRS, self-report) and oxytocin-induced pre-to-post changes in EEG frontal alpha asymmetry. Each point denotes the scores of an individual from the oxytocin group (n= 17). Participants with higher self-reported SRS-motivation scores (lower social motivation) showed a stronger treatment-induced increase in EEG frontal asymmetry (more approach-related motivation).

## Discussion and conclusion

In this study, adopting a randomized, placebo-controlled, double-blind, parallel design, we assessed the effects of a single dose of administered intranasal oxytocin on the approach/avoidance motivational system from a neurophysiological perspective in a context of live gaze social contact. EEG frontal alpha asymmetry was shown to be modulated, upon direct eye contact with a live model, by a single dose of intranasal oxytocin, indicating an increase in approach-related motivational tendencies toward the presented eye contact stimulus. Notably, the treatment effect on frontal asymmetry was most prominently observed in participants with lower self-reported social motivation (higher Motivation subscale scores on the Social Responsiveness Scale; SRS), indicating that participants with lower social motivation benefitted the most from the administered intranasal oxytocin. No treatment-specific changes were identified in terms of gaze behavior toward the eye region of the live model.

### Effect of intranasal oxytocin on frontal alpha asymmetry

While initial studies have related asymmetric frontal alpha activity to emotional valence (i.e. greater left-sided/right-sided activity associated with positive/negative affect or valence), later models suggest that asymmetric frontal alpha activity primarily reflects motivational direction (Harmon-Jones & Gable, 2018). In particular, while in most cases, positive/negative affective situations are associated with approach/avoidance motivation, research has revealed that anger, for example, being an emotional state with a predominantly negative valence, is associated with both approach (relative left-sided frontal activity) and avoidance (relative right-sided frontal activity), and that avoidance is mostly evident when anger is associated with fear or anxiety (Zinner *et al*., 2008; Harmon-Jones & Gable, 2018). Furthermore, it has been shown that differences in the degree of approach motivation can be found while maintaining positive affect constant (Harmon-Jones, *et al*., 2008a).

In the present study, intranasal oxytocin treatment resulted in an increase in left-sided frontal asymmetry scores, while a live model directly gazed at participants, but not when the model sat in front of the participants with eyes closed. Supported by previous research linking EEG relative left-sided frontal asymmetry with approach motivation (Harmon-Jones & Allen, 1997; Sutton & Davidson, 1997; Coan & Allen, 2003; Amodio *et al*., 2008; Gable & Harmon-Jones, 2008; Harmon-Jones, *et al*., 2008a, b; Harmon-Jones *et al*., 2011a, b; Schöne *et al*., 2016;), this result is indicative of an increased approach motivational tendency toward the presented eye contact stimulus.

To our knowledge, this is the first study that directly assesses frontal alpha asymmetry upon oxytocin administration. Rather than studying intranasal oxytocin effects on frontal asymmetry as an outcome measure of a manipulated scenario, Huffmeijer and colleagues (2012) measured frontal asymmetry during rest, adopting this measure as a state and trait factor. They investigated in young females the predictive role of frontal alpha asymmetry, as well as its moderation role on oxytocin and parental love withdrawal, on charity donations. Overall, they found that greater relative left-sided frontal activity (higher frontal alpha asymmetry scores), reflective of higher approach motivation, was associated with larger donations. However, only for participants showing greater relative right-sided frontal activity during baseline (lower scores of asymmetry), associated with avoidance tendencies, was lower parental love withdrawal associated with larger donations after intranasal oxytocin administration. These authors argued that the effects of intranasal oxytocin were only evident on participants with a greater baseline right-sided frontal activity due to the fact that, for those with greater left-sided activity, the approach-related tendencies were already in place prior to treatment. Equally in line with this notion, a study measuring the influence of oxytocin on processing socially relevant cues (Groppe *et al*., 2013), found that women scoring low in sociability had greater effects of oxytocin on a social task. Similarly, increases in relative left-sided frontal asymmetry in our sample were higher for those participants whose scores were also higher in the subscale motivation of the Social Responsiveness Scale, indicating lower motivation towards social contact (e.g., “I don’t join group activities unless I am told to do so”). These findings are in line with the increasing body of evidence showing that oxytocin treatment may be most beneficial for individuals displaying low baseline levels of social proficiency or social approach motivation. Thus, administration of intranasal oxytocin is not likely to induce enhancement of prosocial behaviors when baseline levels of social approach motivation are already high in first place (Bartz *et al*., 2015). For example, in terms of social-cognitive competences, it has also been shown that oxytocin was able to improve empathic accuracy on an emotion recognition task, but only for less-socially proficient men (Bartz *et al*., 2010), but not women (Bartz *et al*., 2019).

Together, these observations highlight the need for thorough characterizations of baseline differences in person-dependent factors, in order to predict the relevance and beneficial effects of intranasal oxytocin treatment regimes. These implications are of particular relevance for the wide variety of neuropsychiatric conditions for which intranasal oxytocin is increasingly considered as a potential treatment (e.g. autism spectrum disorder, social anxiety disorder and depression).

In general, the identified effects of intranasal oxytocin on approach/avoidance related tendencies provide experimental support to the ‘Social Approach/Withdrawal’ hypothesis proposed by Kemp & Guastella (2011), which suggests that oxytocin’s complex role in modulating prosocial behavior results from a down-regulation of withdrawal-related behavior and an up-regulation of approach-related behavior, by inducing a decrease in the experience of social threat (anxiolytic effect) and an increase in the recognition of social reward-driven stimuli (enhancement of social salience), respectively (Kemp & Guastella, 2011). More recently, a more general version of this theory was formulated by Harari-Dahan & Bernstein (2014), namely the ‘General Approach/Avoidance Hypothesis of Oxytocin’ (GAAO), suggesting that oxytocin may not only modulate approach/avoidance motivational tendencies upon social stimuli only, but that the effects may generalize across different types of stimuli when these are emotionally relevant and evocative.

Irrespective of the type of stimulus, it is anticipated that modulations of avoidance-related behaviors are mediated by intranasal oxytocin’s effects on threat-processing networks at the neural level (including the amygdala), whereas modulations of approach-related motivational tendencies are anticipated to be mediated by an up-regulation of reward (dopaminergic) networks (Harari-Dahan & Bernstein, 2014). Initial evidence of a role of oxytocin in modulating amygdala activity, a core hub of the threat-processing network, came from the seminal brain imaging study by Kirsch *et al*. (2005), showing that intranasal oxytocin attenuated amygdala reactivity upon threatening stimuli. Later, Gamer *et al*. (2010) extended this finding by showing that, while intranasal oxytocin reduced amygdala activation upon fearful faces, happy faces tended to elicit stronger activation in certain amygdala subregions after oxytocin administration, thus showing that depending on the valence of the stimuli, and by activating different subregions, the amygdala might facilitate approach motivation for the processing of positive stimuli or reduce avoidance motivation for other negative valenced stimuli. With respect to up-regulation of approach-related motivational tendencies, Shamay-Tsoory & Abu-Akel (2016) proposed in the ‘Social Salience Hypothesis of Oxytocin’ that through its action on dopaminergic reward networks, oxytocin may enhance the salience of stimuli, and that also in this reward-related system, subregions of the amygdala play a central role.

With the current study, we extend the existing neurophysiological evidence supporting a role of intranasal oxytocin in approach/avoidance related tendencies by showing that a single dose of intranasal oxytocin specifically enhances relative left-sided EEG frontal asymmetry, a demonstrated neurophysiological marker of approach-related motivational tendencies. In bringing together our results with the role of the amygdala in approach-related modulations, it is worth mentioning a study by Zotev and colleagues (2016), where EEG and hemodynamic signals, from participants engaged in an emotion regulation task, were simultaneously measured. In this study, participants suffering from major depression disorder were given neurofeedback of their amygdala activity through real-time functional magnetic resonance, while performing an emotion regulation task. Interestingly, while engaged in the neurofeedback task, simultaneously measured EEG revealed that increases in amygdala laterality (being the left amygdala more activated than the right amygdala) correlated with positive EEG asymmetry changes associated with approach motivation (Zotev *et al*., 2016). Thus, it seems that these two measures are both reflective of self-regulation towards approach motivation.

### Effect of intranasal oxytocin on eye gaze behavior

Besides looking at approach motivation from a neurophysiological perspective, we also assessed changes in gaze behavior (fixation count and fixation time towards the eye region of the model) after oxytocin administration. Here, intranasal oxytocin administration did not enhance spontaneous gaze behavior towards the eye region of the observed model’s face. In fact, although non-significantly, both groups (oxytocin and placebo) experienced a decrease in fixation count and fixation time during both model’s eyes open (with direct gaze) and eyes closed condition (see **Table 1**), with no differences between either group or condition. Considering this lack of differences, and that relative left-sided frontal asymmetry only changed in the oxytocin group during eyes open condition, the effect of oxytocin on approach motivation, measured as an increase in relative left-sided frontal asymmetry, cannot be explained from behavioral gazing changes.

Viewing both neutral and emotional faces, some eye-tracking studies have found increases in attention to the eye region (Guastella *et al*., 2008; Gamer *et al*., 2010; Auyeung *et al*., 2015), leading to the idea that such increase mediates other oxytocin-related effects. However, including the present study, not all studies have found increases in visual attention towards the eye region of observed faces after administration of a single dose of intranasal oxytocin. In fact, studies directly testing the effect of oxytocin on different measures of social cognition while taking gaze behavior into account, have countered this notion, showing that oxytocin-related improvements can occur independently of oxytocin effects on eye-gaze patterns (Domes *et al*., 2010; Lischke *et al*., 2012; Hubble *et al*., 2017; Prinsen *et al*., 2018).

In bringing our results together within a motivation framework, we consider it necessary to first stablish a distinction between two motivation concepts: motivational potential and motivational intensity (Brehm & Self, 1989). Whereas motivational potential signals the maximum amount of energy that an organism has available to spend, motivational intensity reflects the actual located energy with respect to an object. Here, social contact is enabled by the live model while directing their gaze at participants, or precluding it by closing their eyes. In both cases, participants are not required to engage in further contact with the live model besides gazing at them. Nonetheless, it is commonly viewed that the first condition, eyes open with direct gaze, is socially engaging, even if only eye contact is taking place. On the contrary, the eyes closed condition can be interpreted by participants as a declaration of no social engagement intention, since the model is seen to refrain from focusing their attention on the other during the whole duration of the trial. It is therefore anticipated that during the eyes open condition, oxytocin exerts an increasing effect of motivational potential, elicited in first place by the live model allowing some degree of social approach, making participants feel more motivated towards the live model, which is reflected by an increase in relative left-sided frontal asymmetry. On the contrary, during the eyes closed condition, the model does not allow any degree of social approach and therefore, the potential motivational approach of participants is very low or null, which is reflected by no changes in relative left-sided frontal asymmetry. With respect to motivation intensity, as Harmon-Jones & Gable (2018) describe it, when someone perceives it impossible to obtain a reward in a certain situation, irrespective of the amount of energy they have available to allocate (motivational potential) to obtain it; motivational intensity (behavior) will be low, reflecting an energy conserving response, given that no effort will grant the reward. In our design, no matter how badly our participants felt an approaching attitude towards the live model, reflected by the increased left-sided frontal asymmetry, the model remained looking at them with the same neutral expression, without engaging in any further contact. Thus, increasing the gazing behavior towards the model would result in no benefit or reward at all, and therefore, it is reasonable that no additional energy is allocated by increasing gazing behavior. Besides subjected to reward, gaze behavior largely depends on relevance and saliency of stimuli. In studies finding an increase in gaze behavior upon oxytocin administration, in general, the task required or at least invited, participants to look at the stimuli. In our study on the other hand, the same face, with the exact same expression was seen by each participant throughout the whole experiment.

## Conclusion

To conclude, a single dose of intranasal oxytocin was shown to increase EEG relative left-sided frontal asymmetry upon direct eye contact with a live model, indicating an increase in approach-related motivational tendencies toward the presented eye contact stimulus. Importantly, the effects were particularly evident in participants with low baseline social approach-related motivational tendencies. Together, these observations add neurophysiological evidence to the hypothesized role of intranasal oxytocin in modulating approach/avoidance related behaviors and suggest that inter-individual variance in person-dependent factors need to be considered in order to evaluate the potential benefit of intranasal oxytocin treatment in trials with neuropsychiatric populations, for whom intranasal oxytocin is increasingly explored as a potential treatment.

## Data Availability

The data set discussed in this manuscript will be made available upon reasonable request.

## Acknowledgements

The authors would like to thank Elisa Maes for her contribution with data collection, as well as to all students and participants collaborating in this study.

## Contributions

Author KA designed the study together with author JP. Author JRS and KA managed the literature searches and analyses. Authors KA, ND and JRS undertook the statistical analysis, and authors JRS and KA wrote the first draft of the manuscript. All authors contributed to and have approved the final manuscript.

## Supplementary Information

**Supplementary Table 1.** Side effects after oxytocin and placebo administration. At the end of the experimental session, participants were asked to report whether they presented any of the listed (or other) side effects and to indicate the severity of the side effect (mild, moderate, or severe). The number of oxytocin participants (out of n=28) or placebo participants (out of n=27) that reported any mild, moderate or severe side effects are listed separately for each side effect. Note that one participant in the placebo group did not fill the side-effect form. A significant group difference (Pearson Chi-square test) was noted for the side effect ‘headache’, indicating that a larger number of oxytocin participants reported a (mild) headache.

|  | Mild |  | Moderate |  | Severe |  | Total |  |  |  |
| --- | --- | --- | --- | --- | --- | --- | --- | --- | --- | --- |
|  | Oxytocin | Placebo | Oxytocin | Placebo | Oxytocin | Placebo | Oxytocin | Placebo | Chi-square | p-value |
| Headache | 6 | 1 | 0 | 0 | 0 | 0 | 6 | 1 | **3.89** | **0.05\*** |
| Drowsiness | 6 | 6 | 6 | 7 | 1 | 0 | 13 | 13 | 0.16 | 0.90 |
| Dizziness | 0 | 0 | 1 | 0 | 0 | 0 | 1 | 0 | 0.98 | 0.32 |
| Fainting | 0 | 0 | 0 | 0 | 0 | 0 | 0 | 0 | 0.00 | 1.00 |
| Changes in heart rate or palpitations | 1 | 2 | 0 | 0 | 0 | 0 | 1 | 2 | 0.39 | 0.53 |
| Shortness of breath | 2 | 0 | 0 | 0 | 0 | 0 | 2 | 0 | 2.00 | 0.16 |
| Fever | 0 | 0 | 0 | 0 | 0 | 0 | 0 | 0 | 0.00 | 1.00 |
| Sore throat | 0 | 0 | 0 | 0 | 0 | 0 | 0 | 0 | 0.00 | 1.00 |
| Dry throat/dry mouth | 3 | 1 | 0 | 1 | 0 | 0 | 3 | 2 | 0.18 | 0.67 |
| Hoarseness | 0 | 0 | 0 | 0 | 0 | 0 | 0 | 0 | 0.00 | 1.00 |
| Coughing | 1 | 0 | 0 | 0 | 0 | 0 | 1 | 0 | 0.98 | 0.32 |
| Coughing up mucus | 1 | 0 | 0 | 1 | 0 | 0 | 1 | 1 | 0.00 | 0.74 |
| Congested nose | 1 | 1 | 0 | 0 | 0 | 0 | 1 | 1 | 0.00 | 0.74 |
| Sneezing | 2 | 0 | 1 | 0 | 0 | 0 | 3 | 0 | 3.06 | 0.12 |
| Nasal irritation | 1 | 2 | 0 | 0 | 0 | 0 | 1 | 2 | 0.39 | 0.49 |
| Runny nose | 8 | 2 | 0 | 1 | 0 | 0 | 8 | 3 | 2.62 | 0.01 |
| Watery eyes | 0 | 2 | 0 | 1 | 0 | 0 | 0 | 3 | 3.29 | 0.11 |
| Nausea and/or vomiting | 0 | 1 | 0 | 0 | 0 | 0 | 0 | 1 | 1.06 | 0.49 |
| Abdominal or stomach pain | 0 | 0 | 0 | 0 | 0 | 0 | 0 | 0 | 0.00 | 1.00 |
| Changes in perception of the tongue | 0 | 0 | 0 | 0 | 0 | 0 | 0 | 0 | 0.00 | 1.00 |
| Burning sensation in nose and/or ears | 0 | 0 | 0 | 0 | 0 | 0 | 0 | 0 | 0.00 | 1.00 |
| Muscle pain/cramps | 0 | 1 | 0 | 0 | 0 | 0 | 0 | 1 | 1.06 | 0.49 |
| Skin rash | 0 | 2 | 0 | 0 | 0 | 0 | 0 | 2 | 2.15 | 0.24 |
| Sweating | 0 | 2 | 1 | 0 | 0 | 0 | 1 | 2 | 0.39 | 0.49 |
| Sensitive to fragrances | 1 | 0 | 0 | 0 | 0 | 0 | 1 | 0 | 0.98 | 0.51 |
| Blurred vision | 1 | 1 | 1 | 0 | 0 | 0 | 2 | 1 | 0.31 | 0.51 |

\* Note that this table has previously been reported in (Daniels *et al.*, 2020). \* $p < .05$ .

**Supplementary Table 2.** Modulation of Profile of Mood States per treatment group. To assess changes in mood, participants completed the 32-item short version of the Profile of Mood States (POMS) questionnaire (Mcnair & Lorr, 1964) at the start (before nasal spray administration) and end of the experimental session (post nasal spray administration) for assessment of transient mood levels in five domains: tension (6 items), depression (8 items), anger (7 items), fatigue (6 items) and vigor (5 items). The POMS comprises emotional adjectives for which subjects need to indicate to what extent the word fits their current mood, using a five-point Likert scale from “not at all” to “very well”. Note that reports of mood states were not recorded for 3 participants (all placebo). For each mood state, the table below lists the mean pre-to-post change scores (± standard error; SE) separately for each treatment group (oxytocin, placebo). T and p (two-tailed) values correspond to between-group differences in pre-to-post change scores. As reported in more detail in Daniels *et al*. (2020), a significant effect of treatment was identified for the mood state ‘tension’, indicating a reduction in reported feelings of tension in the oxytocin group, compared to the placebo group. No significant effects of treatment were identified for the other POMS mood states (depression, anger, fatigue, vigor).

| Profile of Mood State (POMS) | Oxytocin |  | Placebo |  | Between-group difference |  |
| --- | --- | --- | --- | --- | --- | --- |
|  | *N* | *Mean ± SE* | *N* | *Mean ± SE* | *t value* | *p* |
| Tension | 28 | -2.43 ± 0.56 | 25 | -0.76 ± 0.54 | **2.13** | **0.04\*** |
| Anger | 28 | -1.43 ± 0.42 | 25 | -0.88 ± 0.31 | 1.06 | 0.29 |
| Depression | 28 | -1.68 ± 0.44 | 25 | -1.52 ± 0.61 | 0.21 | 0.83 |
| Fatigue | 28 | 0.54 ± 0.48 | 25 | 0.76 ± 0.61 | 0.29 | 0.77 |
| Vigor | 28 | -1.43 ± 0.63 | 25 | -2.32 ± 0.61 | -1.01 | 0.32 |

For POMS tension, anger, depression and fatigue; negative scores indicate pre-to-post improvement. \* $p < .05$ . SE: ± standard error.

